# Common genetic variants are associated with increased likelihood of some co-occurring mental health conditions among autistic individuals

**DOI:** 10.1101/2024.11.11.24317097

**Authors:** Adeniran Okewole, Vincent-Raphael Bourque, Mahmoud Koko, Guillaume Huguet, Anders D Borglum, Jakob Grove, Sebastien Jacquemont, Simon Baron-Cohen, Varun Warrier

## Abstract

**IMPORTANCE:** Autism frequently co-occurs with other mental health conditions. In the general population, these co-occurring mental health conditions are highly heritable and genetically correlated; however, the genetic architecture of co-occurring mental health conditions among autistic individuals is unclear.

**OBJECTIVE:** To investigate the relationship between common and rare genetic variation and co-occurring mental health conditions and latent factors among autistic individuals.

**DESIGN:** Cross-sectional

**SETTING:** The study was conducted with the Simons Foundation Powering Autism Research (SPARK) dataset, V9 release (12 December 2022).

**PARTICIPANTS:** Phenotypic data exploration and factor analyses was conducted in 74,204 autistic individuals, and genetic analyses were conducted in a maximum of 17,582 individuals with genetic data.

**MAIN OUTCOMES AND MEASURES:** Genetic analysis was limited to those probands included in the SPARK iWES1 dataset [n=17,582]. SNP heritability estimates and genetic correlations were computed for three factor scores and six diagnostic categories (attention deficit hyperactivity disorder or ADHD, bipolar disorder, depression, schizophrenia, anxiety disorder and disruptive behaviour disorders or DBD) using bivariate GCTA-GREML and LDSC. Polygenic scores were generated using summary statistics from the most recent genome wide association studies (GWAS) for five traits and six conditions. Associations with factor scores and categorical diagnoses were tested separately for polygenic scores (PGS), *de novo* variants (DNVs) and copy number variants (CNVs).

**RESULTS:** 56% of autistic individuals presented a co-occurring psychiatric condition. Confirmatory factor analysis identified three minimally correlated factors: a behavioural factor, cothymic factor, and a ‘Kraepelin’ or thought disorder factor, with SNP heritabilities ranging from 0.21 (s.e. 0.02) for behavioural to 0.09 (s.e. 0.03) for cothymic factor. Among conditions, moderate and significant SNP heritabilites were observed for ADHD (0.18, s.e. = 0.04) and DBD (0.52, s.e. = 0.08). Moderate positive genetic correlations were found between co-occurring ADHD, DBD, anxiety and the three factors in autistic individuals and corresponding conditions in external population cohorts. PGS for ADHD, depression, and educational attainment were significantly associated with all mental health factors and some of the conditions tested. We found no evidence for an association between common variants for autism, rare CNVs, and DNVs in highly constrained genes with increased likelihood of mental health phenotypes among autistic individuals.

**CONCLUSION AND RELEVANCE:** Among autistic individuals, some mental health conditions and all mental health factors are heritable, but have a distinct genetic architecture from autism itself.

**Key Points:** *Question:* Do genetic variants contribute to co-occurring mental health conditions and latent factors in autistic individuals?

*Findings:* In this cross-sectional study of 17,582 autistic individuals with and without co-occurring conditions, we found significant single nucleotide polymorphism (SNP) heritability for co-occurring ADHD, Disruptive Behaviour Disorders (DBD), and mental-health latent factors. Co-occurring ADHD, DBD, anxiety, and all mental health factors among autistic individuals had moderate genetic correlations with corresponding case-control GWAS. We found no evidence linking common genetic variants linked to autism and rare genetic variants with increased likelihood for mental health conditions among autistic individuals.

*Meaning:* Among autistic individuals, the genetic correlates of co-occurring mental health conditions are distinct from that of autism, suggesting that additional genetic factors contribute to the development of these conditions among autistic individuals.

## 1. Introduction

Autism commonly co-occurs with several mental health conditions, with resultant impairment in quality of life^1,2^. Approximately 70% of autistic children meet the criteria for at least one psychiatric condition^3^, and between 54 - 76% of autistic adults have at least one psychiatric diagnosis^4,5^. Understanding why autistic individuals have higher prevalence of mental health conditions is an important immediate issue to be addressed.

Socio-contextual determinants and adverse life experiences are two significant factors associated with increased mental health conditions among autistic individuals^1^. A third factor is genetics. Autism is heritable^14,15^, and is associated with the additive effects of genetic variants across the allelic frequency spectrum^16–21^. In external population cohorts, mental health conditions are partly heritable and genetically correlated^22–24^. Consequently, it is conceivable that mental health conditions among autistic individuals are associated with genetic variants. However, no study to our knowledge has examined this.

At least two hypotheses can explain the link between genetics and mental health conditions among autistic individuals. Common genetic variants linked to autism are moderately positively genetically correlated with ADHD, depression, self-harm behaviours and modestly with schizophrenia, bipolar disorder and eating disorders^14,25–27^. Similarly, rare genetic variants associated with autism overlap with ADHD^28^ and schizophrenia^29,30^. Consequently, one hypothesis is that co-occurring mental health conditions among autistic individuals are heritable and genetically correlated with genetic variants associated with autism. In other words, variation in genetic predispositions to autism among autistic individuals is associated with their mental health profiles. A second hypothesis is that people are more likely to receive a diagnosis of autism if they have additional mental health conditions and corresponding genetic variants. This would suggest that genetic influences distinct from those linked to autism are associated with elevated mental health conditions among autistic individuals.

Mental health conditions often co-occur^31^, even among autistic individuals, possibly because of shared features across these mental health conditions. Latent factor models have demonstrated transdiagnostic utility in the general population^32^, but their applicability to the autistic population is less well established. Studying the associations between latent mental health factors and genetics may help address potential underdiagnosis of mental health conditions among autistic individuals.

Here, we investigated the genetic correlates (common and rare genetic variants) of co-occurring mental health profiles (both categorical conditions and latent factors) in 17,582 autistic individuals from the Simons Foundation Powering Autism Research (SPARK) cohort^33^. We find significant SNP heritability for some categorical conditions and all latent factors. We do not find an association between common genetic variants for autism or rare genetic variants and mental health conditions. In contrast, we identify moderate to high genetic correlations between some mental health phenotypes and their corresponding phenotypes in the external population cohorts. Finally, within-family based analyses demonstrate direct effects of polygenic scores on some mental health phenotypes without evidence for indirect genetic effects through the familial environment.

## 2. Materials and Methods

### Note on terminology

We use “autistic individuals” in line with preferences of the autistic community. We use preferentially neutral terminology, i.e. “likelihood” or “probability”, when referring to autism and co-occurring conditions. Sex refers to sex assigned at birth.

### 2.1. Description of dataset

Participants in this study included individuals with an autism diagnosis in the Simons Foundation Powering Autism Research (SPARK) dataset^33^) (**Supplementary Figure 1**). Phenotype data was available in the V9 release (12 December 2022) and included 123,444 autistic and non-autistic individuals. Phenotype analysis involved 74,204 autistic individuals for whom core descriptive information was available. Genetic data was obtained from the SPARK iWES v2 (January 2023) release, which included 105,838 autistic and non-autistic individuals. These included 69,758 autistic and non-autistic individuals previously in iWES v1 (WES1-4), who were genotyped using array genotyping. To ensure homogeneity in genotyping across all participants and to allow for joint common and rare variant exploration, genetic analysis was limited to 17,582 participants in WES1 and passed quality control.

### 2.2. Overview of phenotypes, data exploration and confirmatory factor analysis

We focussed on the profiles of mental health conditions in 74,204 autistic individuals (**Supplementary Figure 2**). These include ADHD, schizophrenia, bipolar disorder, depression, anxiety (generalised anxiety and social anxiety combined), and disruptive behaviour disorders (DBD; encompassing oppositional defiant and conduct disorders). The anxiety and DBD phenotypes were created to reflect the fact that they share diagnostic boundaries. We focussed on these six as they clustered into three latent dimensions, are relatively common among autistic individuals, and because there are well powered external GWAS enabling us to study the impact of PGS on these phenotypes in autistic individuals.

For these six phenotypes, we calculated the prevalence in SPARK and compared them with known prevalence among autistic individuals and in the general population, and assessed their co-occurrence using tetrachoric correlation.

Given the correlation among these phenotypes, we assessed if there are underlying latent structures, further details of which are provided in the **Supplementary Methods** and **Supplementary Figure 3.** Confirmatory factor analysis was carried out with the lavaan package (version 0.6.15) in R^34^.

### 2.3. Quality control of genetic data

Genotype data quality control and analyses was restricted to individuals of genetically inferred European ancestries, as provided by the SPARK team. Prior to imputation, we removed variants with less than 95% genotyping rate and those not in Hardy-Weinberg Equilibrium (p < 1E-6). We then excluded individuals with genotyping rate less than 98%, and where family data was available, with excess in Mendelian errors. We additionally excluded individuals with excess heterozygosity (greater than 3 standard deviations from the mean), and whose chromosomal sex did not match their reported sex assigned at birth, as these could indicate technical errors. We imputed participants using the TOPMED imputation panel^35^, using the Michigan Imputation Server^36^ (Phasing using Eagle v2.4^37^ and imputation using Minimac4^38^). After imputation, we retained SNPs with a minor allele frequency > 0.5% and an imputation r^2^ > 0.6. We used liftOver^39^ to convert from GRCh38/hg38 to GRCh37/hg19 build.

Following imputation, plink-format data including 9,698,250 variants for 47,170 participants (autistic and non-autistic) in the WES1-4 component of iWES v2 was obtained. Of this, data for 17,582 autistic individuals were kept for analysis; these had a total genotyping rate of 0.997. Filters for SNP missingness (92% threshold) identified 56,853 SNPs with high missingness, which were excluded. All individuals were within the 0.2 threshold for inbreeding coefficient. As a final step, 1,174 variants were removed due to Hardy-Weinberg exact test (p < 1E-6). This left 9,640,223 SNPs which were used for subsequent analysis.

### 2.4. Generation of genetic principal components

To calculate genetic principal components, we first pruned for linkage disequilibrium (LD) [plink^40^ --indep-pairphase 50 5 0.2], after which SNPs in regions with long range LD were identified and removed. Individual relatedness was calculated using KING ^41^ after which principal components were calculated with PC-AIR in GENESIS^42,43^ which accounts for relatedness among individuals.

### 2.5. SNP heritability and genetic correlations

SNP heritabilities and genetic correlations for factor scores and individual phenotypes were computed using single-component GCTA-GREML^44^ (GREML-SC)and Linkage Disequilibrium Score Regression (LDSC)^45^. GREML analysis used individual-level data with a pruned GRM, while LDSC used summary statistics. Heritability estimates were calculated on both observed and liability scales, with population prevalence estimates obtained from Lai et al., 2019^2^. Genetic correlation among categorical conditions and factors in autistic individuals were calculated using GCTA-GREML. Genetic correlation between the factor scores and diagnostic categories with out-of-sample summary statistics for autism from the non-overlapping iPSYCH dataset^16^, anxiety^46^, ADHD^26^, schizophrenia^47^, bipolar disorder^48^, PTSD^49^, depression^50^,and DBD among individuals with ADHD^51^ were calculated using LDSC. Further details are provided in the **Supplementary Methods**.

### 2.6. GWAS of factor scores and co-occurring mental health conditions

GWAS was performed separately for each factor (using factor scores) and co-occurring mental health condition. This was done using fastGWA-GLMM^52,53^ in the GCTA package version v1.94.1 Linux. FastGWA-GLMM implements a linear mixed effects model for traits in a resource-efficient manner. Each GWAS was performed with a total of 17,573 individuals (from 15,798 families) and for 8,534,720 SNPs with missingness rate < 0.10. Covariates included age at registration (to account for the fact that some mental health conditions are diagnosed only in adolescence or later), sex, and the first 10 genetic principal components. Manhattan and quantile-quantile plots are provided in **Supplementary Figure 4**.

### 2.7. Polygenic scoring

Polygenic scoring was done using PRScs^54^which utilises Bayesian regression framework: inference of posterior effect sizes of SNPs is made by using genome-wide association summary statistics and an LD reference panel which is external to the test dataset. Polygenic scores (PGS) were generated for 11 phenotypes using summary statistics from the cited sources: The iPSYCH-2015 autism GWAS^16^, ADHD^26^, schizophrenia^47^, bipolar disorder^48^, depression^50^, anxiety disorder^46^, neuroticism^55^, P factor^32^, intelligence^56^ and educational attainment^57^, with hair colour^58^ as a negative control. We restricted PGS analyses to 13,290 genetically unrelated autistic individuals identified using GCTA-GREML.

### 2.8. Copy number variant and *de novo* variant data

We generated copy number variant (CNV) data from SNP arrays, and filters were applied as in Huguet et al (2021)^59^. All genes fully encompassed in CNVs were annotated with constraint scores. CNV burden scores were calculated as the count of highly constrained (LOEUF topmost decile)^60^ genes, separately for deletions and duplications (see **Supplementary Methods** for details).

*De novo* variants (DNV) were obtained from trios in SPARK as previously described^61^. DNV analysis was limited to 4,976 autistic individuals with available data. For analysis, protein truncating variants (PTVs) were defined as those with one of these coding consequence: “frameshift_variant”, “start_lost”, “stop_gained”, “splice_donor_variant”, “splice_acceptor_variant” or “stop_lost”. Missense variants were filtered for those with an MPC score ≥ 2. Both PTVs and missense variants were further subset to those with Loss-of-function Observed/Expected Upper-bound Fraction (LOEUF) scores in the topmost decile, and individuals were assigned a binary score for their carrier status. Both PTVs and missense variants were combined to create the category of constrained DNVs.

### 2.9. Regression analyses with PGS, CNV and DNV data

Factor scores were scaled using RobustScaler in ScikitLearn^62^, which scales features using methods that are robust to outliers. Logistic regression analysis within the generalised linear model was performed separately for each phenotype of interest, with PGS, DNVs and CNVs in separate models. In the baseline model, we first tested the association between the phenotypes and age at registration, sex, and cognitive impairment. Next, univariate analyses were conducted to test the association of each genetic variable with each factor and categorical condition separately. This was done over three models: Model 1 with age and the first ten genetic principal components as covariates; Model 2 with sex and sex*genetic variable interaction added (to investigate if the effects differ by sex); and Model 3 with presence/absence of cognitive impairment and its interaction with the genetic variable added (to investigate if the effects differ by co-occurring cognitive impairment). See **Supplementary Methods** for further detail. As a positive control, we also investigated the association between DNVs and CNVs with cognitive impairment.

Thereafter, to assess the combined effect of multiple genetic variables, ten PGS (excluding hair colour) with variance inflation factor not exceeding 4 were included in a multiple PGS model for each factor and categorical condition, with age, sex, cognitive impairment, and the first 10 genetic principal components included as covariates.

In addition, given that for phenotypes such as schizophrenia, bipolar disorder and depression, the younger participants had either not yet reached or were still within the age of risk, Cox proportional hazards regression was also utilised in further sensitivity analyses, with each categorical condition as outcome variable and sex, cognitive impairment, and the first ten genetic principal components included as covariates (see **Supplementary Methods**).

To explore direct and indirect genetic effects (i.e., genetic effects mediated through parental familial effects or nurture), a subset of trios (n= 5,236) for which PGS were available for probands, mothers and fathers were reanalysed in regression models with each factor and phenotype separately as outcome of interest. Analysis was done separately only for PGS with significant effects in the baseline model, with the corresponding PGS for both parents included in each model as previously done^63^. We included age, sex, cognitive impairment and the first 10 genetic principal components derived from the child’s genotypes as covariates.

Across all analyses, multiple testing correction was done using Benjamini-Yekutieli method of false discovery rate.

### 2.10. Ethics statement

The Cambridge University Human Biology Research Ethics Committee provided ethical approval to access and analyse de-identified data from SPARK. The SPARK study is funded by the Simons Foundation and uses a single, central Institutional Review Board (WCG IRB Protocol #20151664). Informed consent was obtained by the SPARK study team from the participant or the caregiver/parent.

## 3. Results

### 3.1. Phenotypic profiles of co-occurring health conditions in autistic individuals

Among autistic individuals in the SPARK cohort (n=74,204; median age at registration = 9 years [IQR = 9 years], 26% female, 56% having a co-occurring psychiatric condition and 16.3% with intellectual disability or cognitive impairment), the prevalence of the six mental health conditions was higher than the general population, but similar to the prevalence previously published in autistic populations (Lai et al, 2019) (**Supplementary Table 1**). After accounting for the age of the participants, these conditions exhibited significant sex differences - with anxiety, depression and bipolar disorder more prevalent among females and ADHD more prevalent among males. With the exception of schizophrenia, all conditions were significantly more prevalent among autistic individuals without co-occurring cognitive impairment (**Supplementary Table 1**). We subsequently investigated the correlation and co-occurrence among the six mental health conditions. We identified correlations ranging from 0.63 (ADHD and DBD, and schizophrenia and bipolar disorder) to 0.19 (ADHD and schizophrenia) (**Figures 1a and b**).

**Figure 1.**
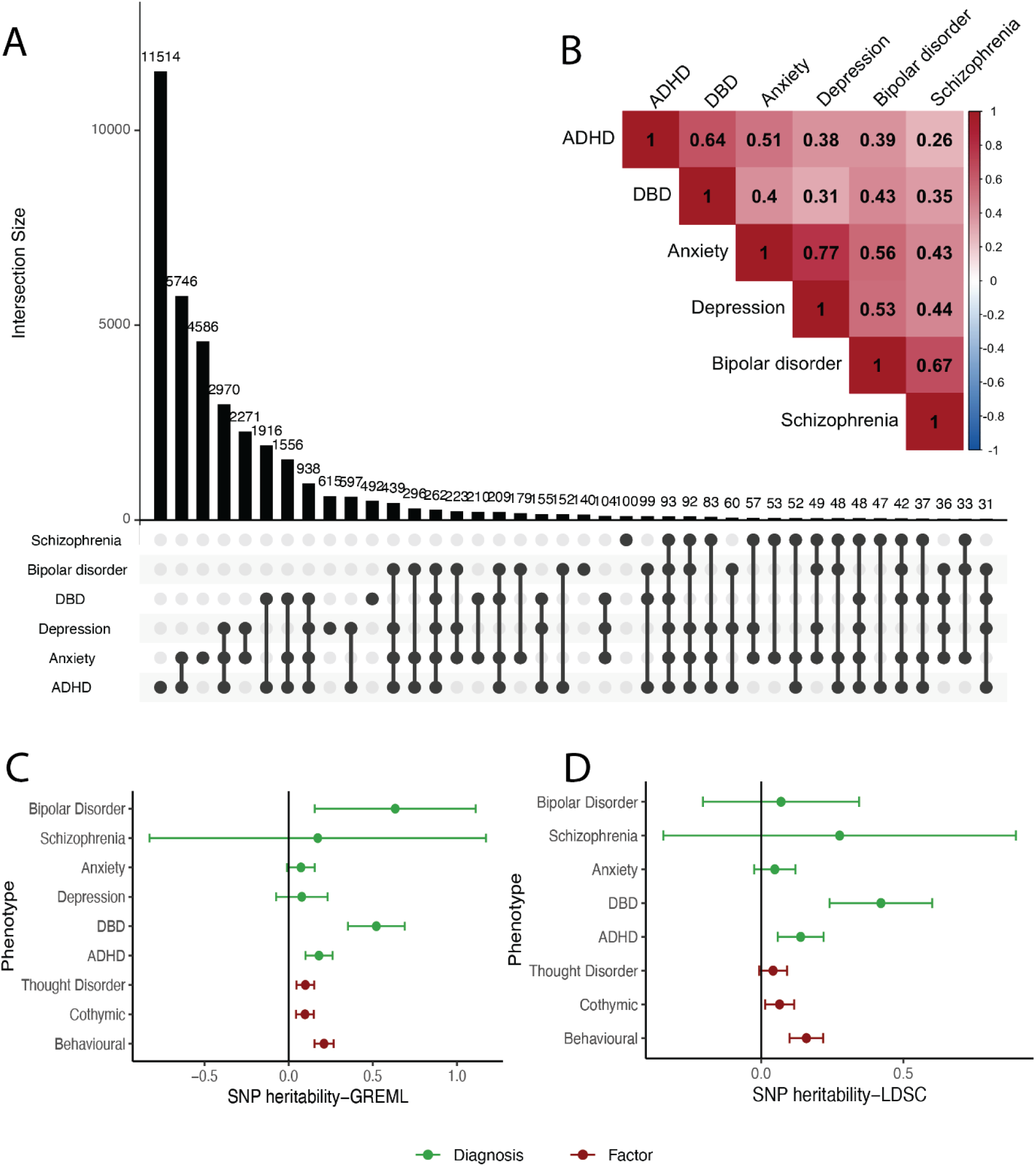
Phenotypic overlap and correlation among the six conditions and SNP-based heritability. A: Upset plot showing overlapping diagnoses among autistic individuals. DBD: disruptive behaviour disorder; ADHD: attention deficit hyperactivity disorder. Sample sizes are: ADHD = 27,573, Anxiety = 20,757, Bipolar disorder = 2,633, DBD = 6,375, Depression = 9,231, Schizophrenia = 988. B: Phenotype correlations of the six co-occurring diagnoses. C: SNP heritability estimates (GCTA-GREML) of the diagnoses and factors. Point estimates indicate heritability, and whiskers indicate 95% confidence intervals. D: SNP heritability estimates (LDSC) of the diagnoses and factors. Point estimates indicate heritability, and whiskers indicate 95% confidence intervals.

Given this correlation, we conducted confirmatory factor analyses to identify latent factors. The best performing model (**Supplementary Table 2** and **Supplementary Figure 2**) included three factors - a behavioural factor (including ADHD and disruptive behaviour disorders), a cothymic factor (comprising anxiety and depression), and a ‘Kraepelin’ or thought disorder factor which grouped schizophrenia and bipolar disorder together (CFI = 0.972, TLI = 0.929, RMSEA = 0.05).

### 3.2. Common variant genetic architecture of co-occurring mental health conditions

To understand if common genetic variants are associated with co-occurring mental health conditions and factors, we calculated SNP heritabilities on the liability scale using GREML-SC and LDSC (**Supplementary Table 3 and Figure 1C and 1D**). Both methods identified moderate and significant SNP heritabilities for behavioural conditions (ADHD and DBD) and the behavioural factor. In addition, we identified significant but low SNP heritabilities for cothymic and thought disorder factors. The heritability of the other phenotypes failed to reach statistical significance, potentially due to the low statistical power and underdiagnosis of younger autistic individuals.

We further investigated the genetic correlation among these phenotypes. We identified high genetic correlations among the 3 factors (**Supplementary Table 4**). Using GREML-SC, among categorical conditions, we identified high and significant genetic correlation among DBD, anxiety, and ADHD, and high genetic correlation between bipolar disorder and both DBD and anxiety. Estimates of these genetic correlations were consistent between GREML-SC and LDSC.

Subsequently, we calculated genetic correlations between the co-occurring factors and conditions, and seven external GWAS of psychiatric conditions, including autism. The external GWAS for ADHD, DBD, PTSD, and depression had significant and moderate genetic correlations with the behavioural factor, cothymic factor, ADHD and DBD in autistic individuals (**Supplementary Table 5 and Figure 2**).

**Figure 2.**
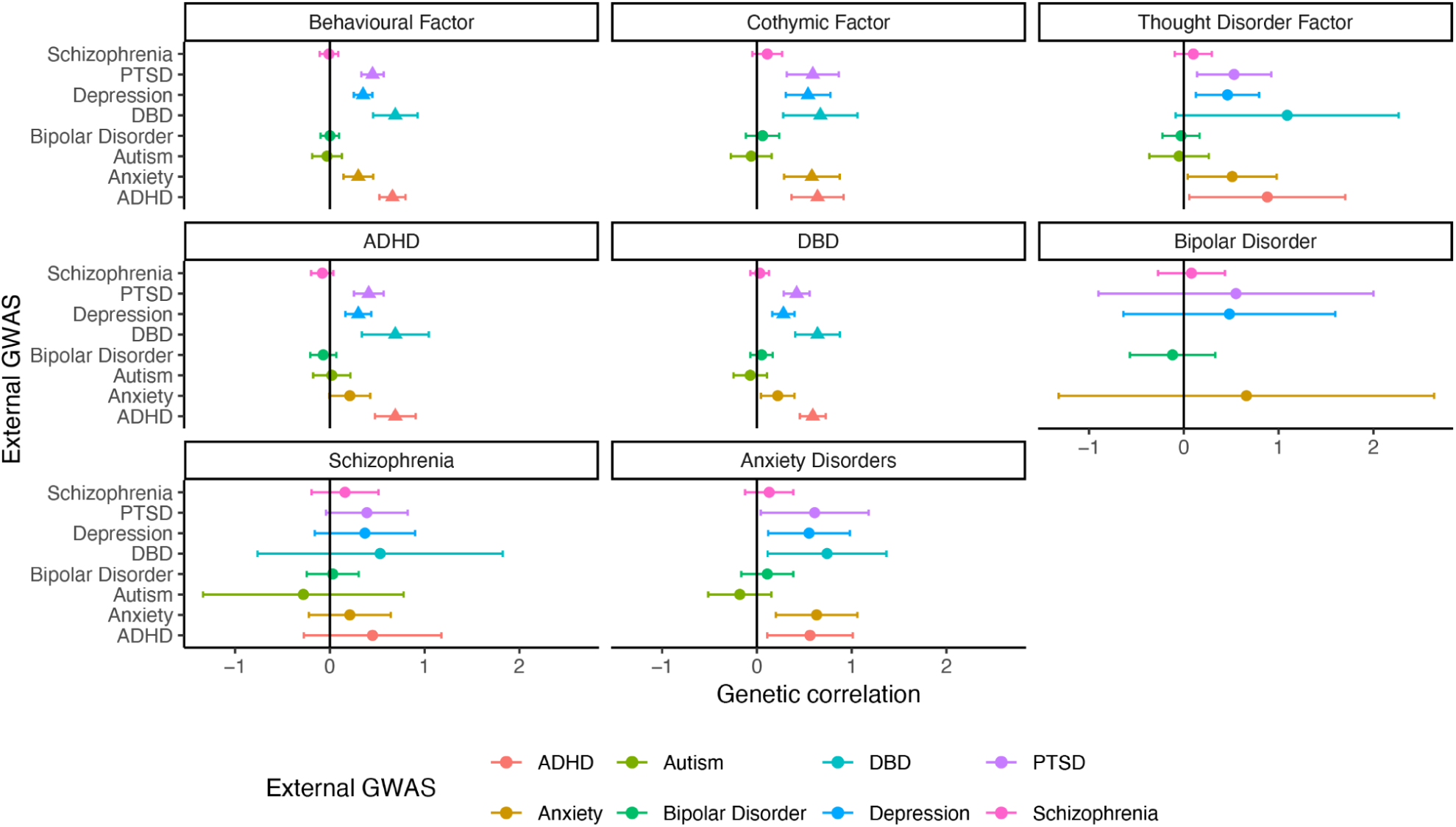
Genetic correlations between mental health phenotypes in autistic individuals and external population cohorts (LDSC). Point estimates indicate genetic correlation estimates, and whiskers indicate 95% confidence intervals. Triangles indicate significant genetic correlations after Bejamini-Yekutieli (BY) multiple testing correction.

### 3.3. Genetic associations of factor scores and co-occurring mental health conditions

To further investigate the link between genetic variants and co-occurring mental health phenotypes, we ran regression between genetic variants (CNV scores, DNVs, and PGS) and the mental health phenotypes. In univariate models (Model 1), PGS for ADHD, depression, and educational attainment were significantly associated with all phenotypes in the expected direction (**Supplementary Tables 6** and **8, Figure 3**). We obtained consistent results when accounting for cognitive impairment (Model 3), and consistent results for the three factors, DBD, and ADHD when accounting for sex (Model 2). In addition, we observed significant associations between PGS for anxiety and IQ for several of these phenotypes. Our control PGS (hair colour) was not associated with any phenotypes.

**Figure 3.**
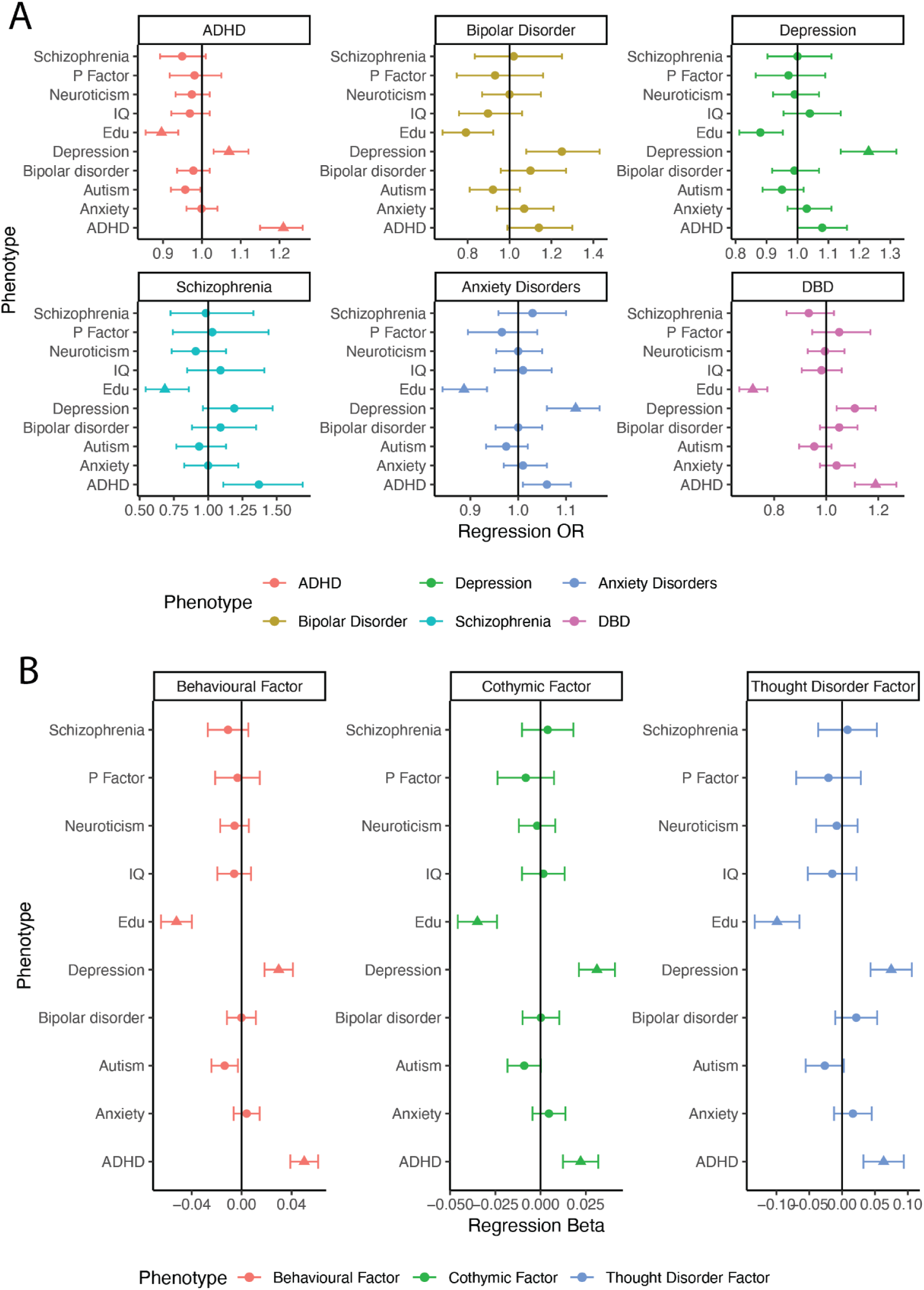
Association between PGS and mental health phenotypes among autistic individuals. A: Regression with phenotypic categorical diagnoses. DBD: disruptive behaviour disorder; ADHD: attention deficit hyperactivity disorder; DEP: depression; BIPD: bipolar disorder; ANX: anxiety disorder; SCZ: schizophrenia. B: Regression models with robustly scaled factor scores and 10 polygenic scores (PGS). Edu: educational attainment PGS. For both plots, point estimates indicate regression coefficients and whiskers indicate 95% confidence intervals. Triangles represent PGS with significant association after Bejamini-Yekutieli (BY) multiple testing correction.

While rare genetic variants significantly impacted the likelihood of cognitive impairment as expected (**Figure 4** and **Supplementary Figure 5**), with the largest effects observed for *de novo* PTV variants followed by deletions, *de novo* missense variants, and duplications, we identified no significant association between any of these rare genetic variants and any of the mental health phenotypes. We obtained largely consistent results when using Cox proportional hazard regression (**Supplementary Table 9**), and identified no significant interaction with sex or cognitive impairment (**Supplementary Tables 6** and **8**).

**Figure 4:**
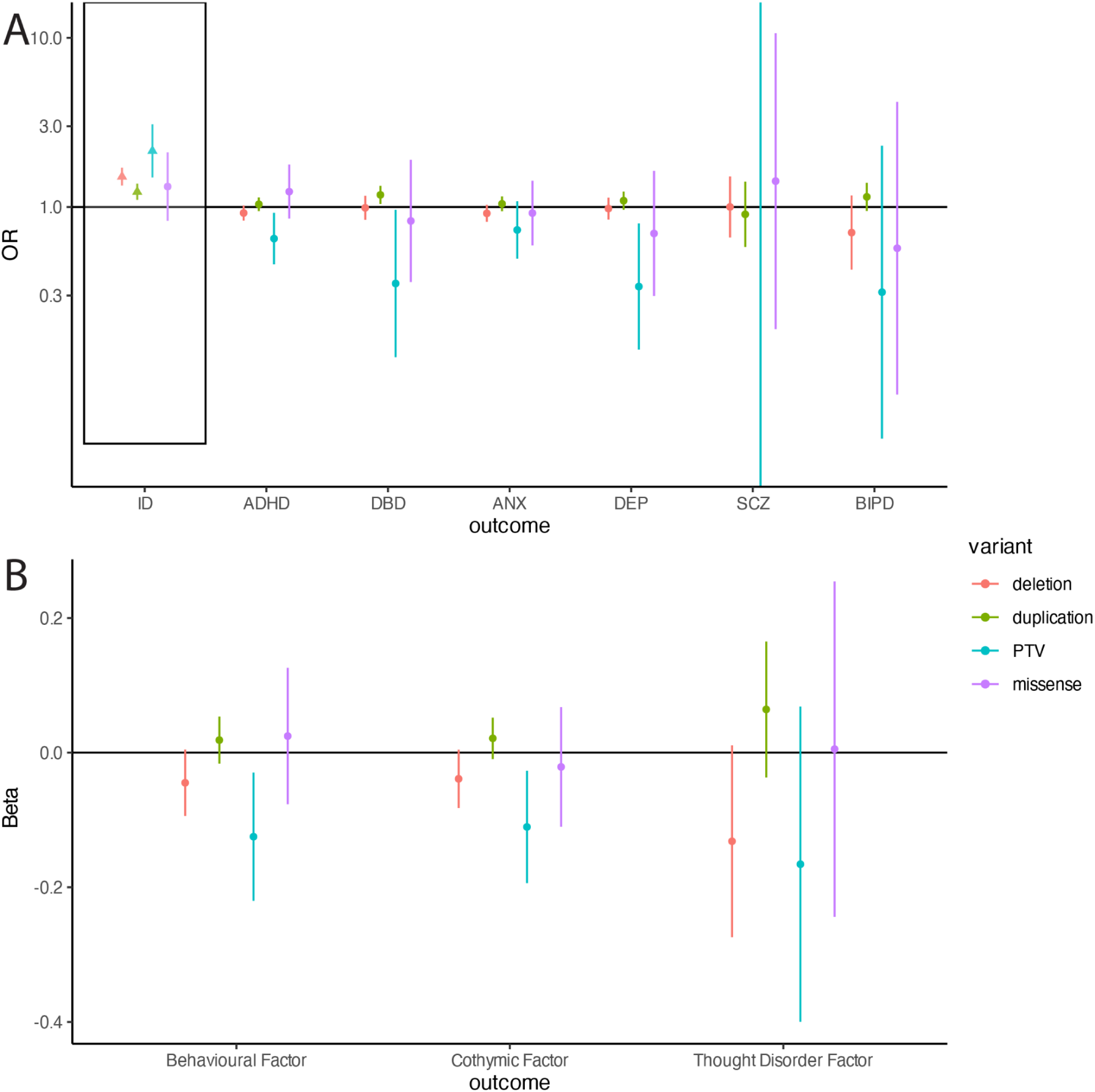
Association between rare-variants and mental health phenotypes among autistic individuals. A: Regression models for intellectual disability and categorical diagnoses. B: Regression models for robustly scaled factor scores. ID: intellectual disability. P_adj: adjusted p values; ns: not significant. PTV: protein truncating variants. For both plots, point estimates indicate regression coefficients and whiskers indicate 95% confidence intervals. Triangles represent PGS with significant association after Bejamini-Yekutieli (BY) multiple testing correction. ID = intellectual disability, ANX = anxiety, DEP = depression, SCZ = schizophrenia, BIPD = Bipolar disorder

We followed up the univariate analyses with multivariate analyses to account for the correlation among polygenic scores. In these, we simultaneously regressed ten PGS against the outcome phenotypes. In these conditional analyses, PGS for ADHD, depression, and EA were associated in the expected direction with all factors (**Supplementary Table 7**) and the majority of the mental health conditions (**Supplementary Table 10**). Yet again, we obtained consistent results when using Cox proportional-hazards regression.

We further wondered if the association between PGS for EA, ADHD and depression represent direct genetic effects of the child, or indirect genetic effects of parental nurture. We tested this using models of PGS association including proband and parent PGS for each phenotype (**Supplementary Tables 11** and **12,** and **Figure 5**). We found no evidence of indirect effects - there was no significant association between either paternal or maternal PGS and any of the phenotypes tested. However, we found significant association between the autistic proband’s depression and EA PGS and cothymic and behavioural factors even after conditioning on parental PGS. In addition, the autistic proband’s ADHD PGS was significantly associated with the behavioural factor, and EA PGS with DBD. All associations were in the expected direction.

**Figure 5:**
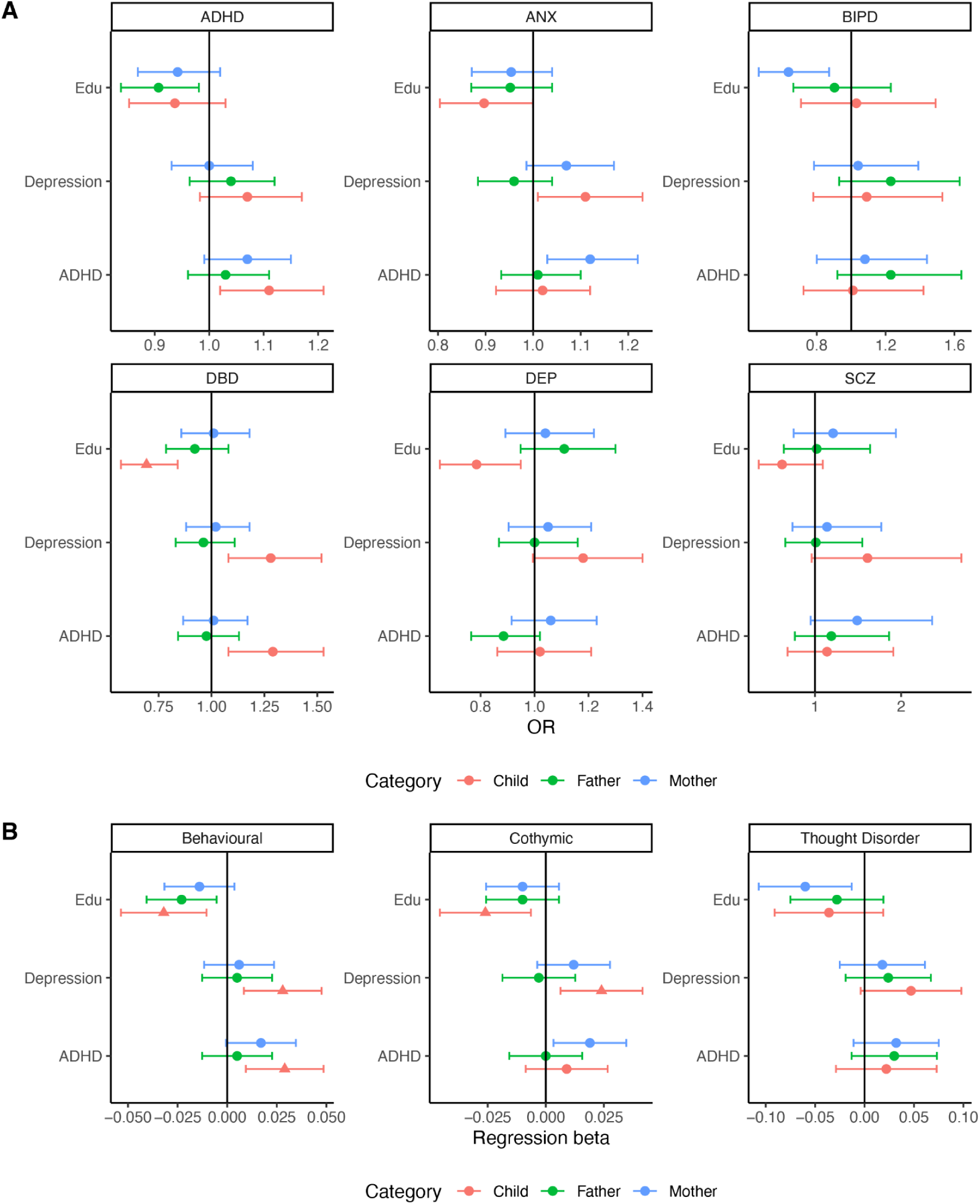
Family (trios) models with PGS. A: Regression models with categorical diagnoses. B: Regression models with robustly scaled factor scores. For both plots, point estimates indicate regression coefficients and whiskers indicate 95% confidence intervals. Triangles represent PGS with significant association after Bejamini-Yekutieli (BY) multiple testing correction. PGS from external GWAS provided on the y-axis. ANX = anxiety, DEP = depression, SCZ = schizophrenia, BIPD = Bipolar disorder

## 4. Discussion

In this study, we conducted the first large-scale genetic investigation of co-occurring mental health phenotypes among autistic individuals from the SPARK cohort. The prevalence of the studied mental health conditions in autistic individuals approximated what has been reported in literature^2^, albeit with deviations for schizophrenia (lower prevalence) and ADHD (higher prevalence). This may reflect the demographic profile of participants in the SPARK cohort.

We highlight five key results. First, we demonstrate that ADHD, DBD, and all mental health factors have modest and significant SNP heritabilities, suggesting that these emerge from complex sociobiological mechanisms. Although other conditions tested did not have significant SNP heritabilities, this may be because of low statistical power (e.g., schizophrenia), and because the majority of autistic individuals are young and may not have reached an appropriate age for diagnosis. Second, we identify moderate to high genetic correlations between co-occurring ADHD, DBD, anxiety and all three mental health factors among autistic individuals and the corresponding conditions in external population cohorts. This suggests similar genetic variants are associated with mental health phenotypes both among autistic individuals and external cohorts. Third, neither PGS analyses nor genetic correlation studies identify shared genetics between autism and co-occurring mental health phenotypes specifically among autistic individuals. There is a positive and moderate genetic correlation between autism and other mental health conditions in the external population cohorts, suggesting that increased polygenic propensity for autism is associated with higher likelihood of mental health conditions. Taken together, this suggests that co-occurring mental health conditions in autistic individuals arise from a combination of their elevated autism polygenic propensity and additional mental health-specific genetic factors in addition to other socio-contextual factors. Fourth, within family analyses indicate that the effects of the PGS on some mental health phenotypes are direct (i.e., effects of inherited genetic variation on the child’s phenotype) and we find no evidence for indirect genetic effects (i.e., parental genetics influencing the autistic child’s mental health through the familial environment). Fifth, and finally, we find no evidence for the role of rare genetic variants on increasing the likelihood for co-occurring mental health conditions among autistic individuals, despite an association between rare variants and mental health conditions in external population cohorts^28,64–66^. This pattern may reflect collider bias, limited statistical power, phenotypic heterogeneity between autistic individuals with and without rare variants^16^, or diagnostic overshadowing of mental health conditions^67,68^ in autistic individuals with rare variants who may have complex support needs. The finding of heritable latent factors suggests the utility of a biaxial system of diagnosis, with dimensional latent factors on one axis and conventional categorical diagnosis on another.

For oppositional defiant and conduct disorder, which together form the kernel of disruptive behaviour disorders in ICD-11^69^, there was no comparable SNP heritability in external population cohorts. Notably, the GREML-SC and LDSC based SNP heritability for DBD was high: 0.52 (se: 0.08) in line with a previous study on the SNP heritability for DBD among individuals with ADHD (h^2^SNP = 0.34; SE = 0.05)^51^. The finding of significant genetic correlation between disruptive behaviour disorders in the SPARK and iPSYCH cohorts suggests the two constructs are consistent. As observed by Sesso et al (2020)^70^, autism and disruptive behaviour disorders represent distinct phenotypes among individuals with ADHD. Given the high heritability of DBD among autistic individuals, it may be possible that autistic individuals with co-occurring DBD represent a distinct subgroup.

PGS for ADHD and depression were significantly associated with several phenotypes. However, among all the PGS, the most consistent association with all the co-occurring phenotypes was with educational attainment, reflecting the shared genetics between educational attainment and a wide number of mental health conditions in population cohorts^71^.

This study has a few limitations. First, it was conducted with only one dataset, which may limit its generalisability. Future work will seek to replicate the study in other comparable datasets. Second, there was also a modest number of participants with some conditions - especially schizophrenia and bipolar disorder - which may have accounted for the lack of precision of some estimates. The low sample size also meant that we had limited statistical power to detect more modest effects for DNVs and in trio-based analyses of PGS. Furthermore, mental health conditions may be underdiagnosed in carriers of DNVs who often have complex support needs. Third, other factors which may have influenced the heritability and genetic correlation estimates include sample overlap, assortative mating and collider bias. Specifically, for depression, we were unable to estimate genetic correlation using LDSC both among autistic individuals and with the external population due to the low SNP heritability. Fourth, we were able to investigate the genetic correlates of mental health conditions only among individuals of genetically inferred European ancestries, and the generalisability of these findings to other population groups is unknown. Furthermore, for depression, we were unable to estimate genetic correlation using LDSC both among autistic individuals and with the external population due to the low SNP heritability.

## Data Availability

All data produced in the present study are available upon reasonable request to the authors

## ACKNOWLEDGEMENTS

This study was conducted as part of AO’s doctoral programme at the University of Cambridge, funded by the Cambridge Trust and a Trinity-Henry Barlow studentship. SBC received funding from the Wellcome Trust 214322\Z\18\Z. For the purpose of Open Access, the author has applied a CC BY public copyright licence to any Author Accepted Manuscript version arising from this submission. SBC also received funding from the Innovative Medicines Initiative 2 Joint Undertaking under grant agreement No 777394 for the project AIMS-2-TRIALS. This Joint Undertaking receives support from the European Union’s Horizon 2020 research and innovation programme and EFPIA and AUTISM SPEAKS, Autistica, SFARI. SBC also received funding from the Autism Centre of Excellence (ACE) at Cambridge, SFARI, the Templeton World Charitable Fund, the MRC, and the NIHR Cambridge Biomedical Research Centre. The research was supported by the National Institute for Health Research (NIHR) Applied Research Collaboration East of England. Any views expressed are those of the author(s) and not necessarily those of the funder. The funders had no role in the design of the study; in the collection, analyses, or interpretation of data; in the writing of the manuscript, or in the decision to publish the results. Any views expressed are those of the author(s) and not necessarily those of the funders (including IHI-JU2, the NIHR and the Department of Health and Social Care).

## Notes

### Author Declarations

We obtained ethics approval from the Cambridge Human Biology Research Ethics Committee to analyse pseudonymised human genetic data.

